# Health phenome of Parkinson’s patients reveals prominent mood-sleep cluster

**DOI:** 10.1101/2022.02.01.22270276

**Authors:** Abby L. Olsen, Joseph J. Locascio, Idil Tuncali, Nada Laroussi, Elena Abatzis, Polina Kamenskaya, Yuliya Kuras, Tom Yi, Aleksandar Videnovic, Michael T. Hayes, Gary P. H. Ho, Jordan Paulson, Vikram Khurana, Todd M. Herrington, Bradley T. Hyman, Dennis J. Selkoe, John H. Growdon, Stephen N. Gomperts, Trond Riise, Michael A. Schwarzschild, Albert Y. Hung, Anne-Marie Wills, Clemens R. Scherzer

## Abstract

**Introduction:** Parkinson’s disease (PD) patients have multiple co-morbidities, exposures, and medications. Our objective is to evaluate the complexity of features associated with PD in a study of 933 cases and 291 controls in the Harvard Biomarkers Study.

**Methods:** The primary analysis evaluated 64 health features for associations with PD using logistic regression adjusting for age and sex. We adjusted for multiple testing using the false discovery rate (FDR) with ≤ 0.05 indicating statistical significance. Exploratory analyses examined feature correlation clusters.

**Results:** Depression (OR = 3.11, 95% CI 2.1 to 4.71), anxiety (OR = 3.31, 95% CI 2.01-5.75), sleep apnea (OR 2.58, 95% CI 1.47-4.92), and restless leg syndrome (RLS; OR 4.12, 95% CI 1.81-12.1) were more common in patients with Parkinson’s than in controls. The prevalence of depression, anxiety, sleep apnea, and RLS were correlated, and these diseases formed part of a larger cluster of mood and sleep traits linked to PD. Exposures to pesticides (OR 1.87, 95% CI 1.37-2.6), head trauma (OR 2.33, 95% CI 1.51-3.73), and smoking (OR 0.57, 95% CI 0.43-0.75) were also significantly associated with PD. Vitamin D3 (OR 2.18, 95% CI 1.4-3.45) and coenzyme Q10 use (OR 2.98, 95% CI 1.89-4.92) were more common in PD. Cumulatively, 43% of PD patients had at least one psychiatric or sleep disorder, compared to 21% of controls.

**Conclusions:** 43% of PD patients in an academic medical center have depression, anxiety, or disordered sleep. This syndromic cluster of mood and sleep traits may be pathophysiologically linked and clinically important.

## Introduction

Parkinson’s disease (PD) is likely to arise through a complex interplay of genetic^1^ and environmental factors^2^. With exceptions^3,4^, most epidemiologic studies have examined one or a few variables at a time. In the real world, patients are much more complex, with phenotypic diversity, varied comorbidities, and polypharmacy. Here we begin to characterize the picture of clinical, pharmacological, and environmental traits linked to patients with PD.

The Harvard Biomarkers Study (HBS; now Yale Harvard Biomarkers Study (YHBS)) includes an extensive questionnaire regarding past medical history, medication and supplement use history, social history, and environmental exposures. This includes data on some previously reported putative risk or protective factors, though not all. We perform age- and sex-adjusted logistic regression to determine which of these health variables are positively or negatively associated with PD. Importantly, these are correlative association and not indicators of causality.

We identify 8 variables positively associated with PD and one negatively associated, including a cluster of co-occurring sleep and mood disorders.

## Methods

### Harvard Biomarkers Study

The Harvard Biomarkers Study (HBS) is a case-control study including thousands of patients with neurodegenerative diseases as well as healthy controls. Informed consent was obtained for all participants. The study protocol was approved by the institutional review board of Mass General Brigham.

This analysis included 1,224 total subjects: 933 with PD and 291 healthy controls based on data availability. Healthy controls consisted chiefly of spouses, friends, and non-blood relatives, who accompanied PD patients to office visits. PD diagnosis was made by a board-certified neurologist with movement disorders fellowship training. Subjects completed a detailed questionnaire (Supplemental Table 1). Each item in the questionnaire is phrased as a binary yes/no question. Data from the enrollment visit were analyzed. At the time of enrollment, average disease duration was 3.8 years (Table 1).

**Table 1.** Clinical characteristics. Statistical differences in demographic data (age, sex, and race) between the cases and controls were determined using Satterthwaite t-test, Chi-square test, and Fisher’s exact test, as appropriate. UPDRS, Unified Parkinson’s Disease Rating Scale. MMSE, Mini-Mental State Exam.

| **Table 1. Clinical characteristics** |  |  |  |
| --- | --- | --- | --- |
| **Characteristic** | **Healthy controls** | **Parkinson's disease** | **p-value** |
| Sex |  |  | <0.0001 |
| Male | 125 (43%) | 620 (66.5%) |  |
| Female | 166 (57%) | 312 (33.5%) |  |
| Age at enrollment (years) | 63.5 ± 11.7 | 66.1 ± 9.9 | 0.0006 |
| Race |  |  |  |
| Asian | 2 (0.7%) | 9 (0.7%) | 1.000 |
| Black/African American | 13 (4.9%) | 9 (0.7%) | <0.0001 |
| Hawaiian/Pacific | 1 (0.3%) | 0 | 0.19 |
| More than one race | 3 (1%) | 3 (0.2%) | 0.09 |
| Unknown | 4 (1.4%) | 38 (3.1%) | 0.12 |
| European | 268 (92.1%) | 1166 (95.2%) | 0.04 |
| Ethnicity |  |  |  |
| Hispanic | 8 (2.7%) | 17 (1.4%) | 0.12 |
| Non-hispanic | 277 (95.2%) | 1168 (95.3%) | 0.88 |
| Unknown | 6 (2.1%) | 40 (3.3%) | 0.34 |
| Education (years, S.D.) | 15.0 ± 1.77 | 15.2 ± 1.76 | 0.16 |
| Age at onset |  | 62.3 ± 10.4 |  |
| Disease duration |  | 3.8 ± 4.4 |  |
| Hoehn and Yahr stage |  | 2.1 ± 0.64 |  |
| UPDRS motor subscale |  | 19.8 ± 10.2 |  |
| UPDRS total |  | 33.2 ± 15.1 |  |
| MMSE | 28.99 ± 1.26 | 28.21 ± 2.30 | <0.0001 |

Statistical differences in demographic data (age, sex, and race) between cases and controls were determined using a Satterthwaite t-test, Chi-square test, or Fisher’s exact test, as appropriate (Table 1). Included in the table are the mean Unified Parkinson’s Disease Rating Scale (UPDRS) and the Mini-Mental State Exam (MMSE) scores.

### Logistic regression

We tested for association between 64 clinical variables (Figure 1, Supplemental Figure 2) and PD using age- and sex-adjusted logistic regression analysis in SAS version 9.4. The diagnostic group (PD or healthy control) was the dependent variable. Each of the 64 clinical variables was run as a separate logistic regression analysis, and subjects who were missing values on any of the 64 variables were excluded from every analysis. After excluding subjects with missing values, 494 PD and 142 healthy controls were available for this analysis.

**Figure 1:**
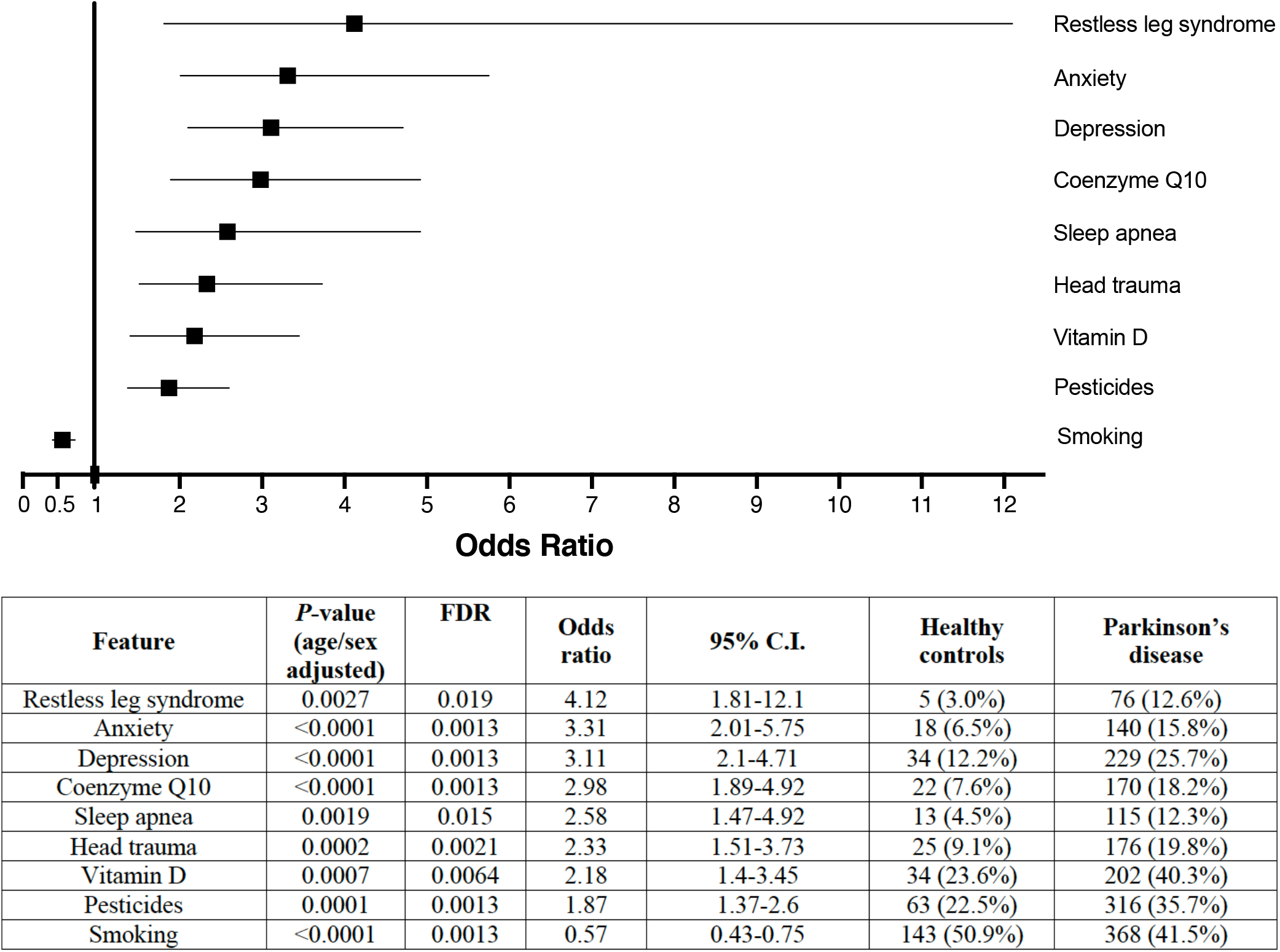
Results of logistic regression. Depression, anxiety, restless leg syndrome, sleep apnea, vitamin D supplementation, coenzyme Q-10 supplementation, head trauma, and exposure to pesticides were over-represented in PD. Smoking was inversely associated with PD. Odds ratio with 95% confidence limit for each variable is shown. In the corresponding table, *P*-value, odds ratio, 95% confidence limit, and prevalence in HC versus PD are shown. Each variable is controlled for sex and age.

### Variable correlation

To determine which predictor variables are statistically similar, we subjected the Pearson correlation matrix of the entire set of 64 variables to a hierarchical clustering algorithm (employing the Corrplot package in R, version 1.1.423 and the graphical software GraphPad Prism, version 8.43).

## Results

Cases averaged three years older than controls (mean age 66 years for PD, 63.5 years for healthy controls, p = 0.0006) and were more likely to be male (66.5% vs 43%, p <0.0001) (Table 1). We thus adjusted our logistic regression for age and sex. 64 features were analyzed for association with PD (Supplemental Table 1, Supplemental Figure 2). Nine features reached statistically significant associations with PD after adjustment for multiple testing using the false discovery rate (FDR) (Figure 1, Supplemental Figure 1). Anxiety (OR = 3.31, 95% CI 2.01-5.75), depression (OR = 3.11, 95% CI 2.1-4.71), sleep apnea (OR 2.58, 95% CI 1.47-4.92), and restless leg syndrome (OR 4.12, 95% CI 1.81-12.1) were more common in PD than in controls adjusting for age and sex with FDR ≤ 0.05. Exposures to pesticides (OR 1.87, 95% CI 1.37-2.6), head trauma (OR 2.33, 95% CI 1.51-3.73), and smoking (OR 0.57, 95% CI 0.43-0.75) were associated with the disease. Vitamin supplementation with cholecalciferol (OR 2.18, 95% CI 1.4-3.45) and coenzyme Q10 (OR 2.98, 95% CI 1.89-4.92) were more commonly used by patients than controls.

Five additional variables had *suggestive* associations (nominal p <0.05) but did not survive correction for multiple testing, including positive associations with ibuprofen, buproprion, and antiemetics, and inverse associations with ezetimibe and multivitamins (Supplemental Table 2).

To examine whether some of these features are correlated and thus may tag the same underlying trait we clustered the pair-wise correlation structure between the 64 variables. The resulting Pearson correlation matrix is shown in Figure 2A. Hierarchical clustering of the correlation coefficients revealed three correlated feature clusters (Figure 2B).

**Figure 2:**
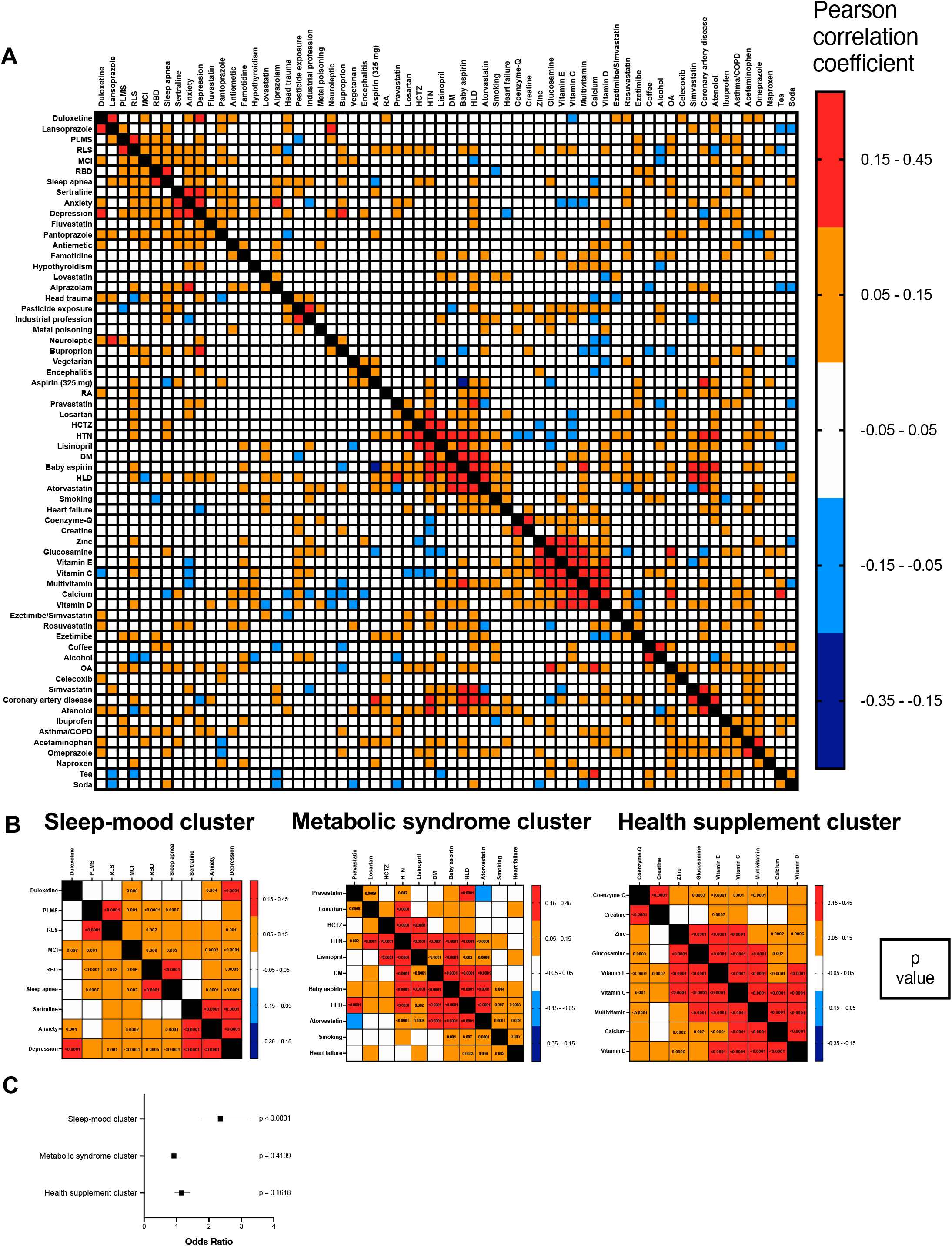
Heat map demonstrating correlations between the individual clinical and environmental variables. A. Pearson correlation matrix. B. Several sets of variables are positively correlated, forming clusters of biologically related phenomena. The largest clusters in our dataset include a linked sleep and psychiatric factor cluster, a metabolic syndrome cluster, and a vitamin use cluster. P-values indicating the significance of the correlation are shown in the boxes. C. Of the 3 clusters, only the mood-sleep cluster is significantly associated with PD.

*Cluster 1* represents nine correlated *mood and sleep traits*. Within this cluster, depression was the variable with the greatest degree of correlation (based on correlation coefficients and p-values) to all other variables in the cluster. *Cluster 2* represents *metabolic syndrome traits*. Within this cluster, hypertension was the variable with the greatest degree of correlation to all other variables. *Cluster 3* represents nine correlated *health supplements*. The high degree of correlation between various vitamin supplements is consistent with our clinical impression that patients who take vitamins are likely to take multiple vitamins.

To determine if the 3 clusters were themselves associated with PD, we repeated the sex and age adjusted logistic regression after combining the variables within each cluster into a single factor score. A factor score for each person was defined as the proportion of the variables within that factor that a person possessed. The mood-sleep factor score (cluster 1) was significantly associated with PD (Figure 2C), while the other two clusters were not associated with PD.

Cumulatively, 42% (396 of 933) of patients had at least one psychiatric or sleep disorder, compared to 21% (61 of 291) of healthy controls. Among the variables in the mood-sleep cluster, depression was the single most prevalent diagnosis in PD patients, with 24.5% of PD patients (229 of 933) having depression, compared to 11.7% of controls (34 of 291). Further, 20% of PD patients (187 of 933) had multiple mood-sleep cluster diagnoses, in contrast to only 5.5% of healthy controls (16 of 290) (Supplemental Figure 2A). Among the PD patients with multiple diagnoses, a large majority (81%, 152 of 187) had depression as one of the diagnoses (Supplemental Figure 2B).

## Discussion

This study represents a multi-dimensional and data-rich view of clinical associations with PD (Supplemental Figure 3) using the deeply characterized HBS cohort^5^. As this is a case-control study, the results should not be interpreted as causal. Our cross-sectional design cannot distinguish exposures and comorbidities that may contribute to PD risk from those that arise as a consequence of a PD diagnosis or its prodrome. That is, while some associations may arise because a variable affects risk of PD, others almost certainly arise because of having a diagnosis of PD affects risk of the variable. For example, our prior work has identified vitamin D3 deficiency in the HBS cohort^6^, which has likely resulted in participants increasing their vitamin D3 intake. Further, because the HBS questionnaire captures each comorbidity as a binary yes/no at the time of enrollment rather than the age or date of onset, we are unable to establish whether these diagnoses preceded or followed the onset of PD motor symptoms in individual participants. Future work incorporating onset-timing data relative to PD diagnosis, or longitudinal designs following at-risk individuals prospectively, will be needed to clarify these temporal relationships.

One striking finding in our analysis was the strong prevalence of mood and sleep disorders in our PD population. The interplay between mood disorders and PD is complex, with some evidence suggesting that these disorders represent either pre-motor or early non-motor co-comorbidities. Similarly, sleep disorders are prevalent in PD and may arise either prior to or after the onset of motor symptoms. REM behavior disorder (RBD) may represent a prodromal state of PD, and there is growing interest in conducting clinical trials in patients with isolated RBD.

Noradrenergic signaling plays a critical role in PD, as well as in the regulation of mood and sleep, suggesting shared pathobiology^7^. In PD, degeneration of locus coeruleus noradrenergic neurons often precedes dopaminergic loss, contributing to non-motor symptoms such as depression, anxiety, cognitive impairment, and sleep disturbances. Reduced brain norepinephrine disrupts modulation of cortical and limbic circuits, which are essential for emotional regulation. Dysregulation of this system can therefore lead to mood disorders, insomnia, fragmented sleep, REM sleep behavior disorder, and excessive daytime sleepiness— features commonly observed in PD. Together, these findings raise the hypothesis that noradrenergic dysfunction may contribute to the co-occurrence of neurodegeneration, affective symptoms, and sleep abnormalities in PD. However, our clustering analysis identifies statistical co-occurrence among depression, anxiety, RLS, and sleep apnea; it does not test or establish a shared biological mechanism.

Our study reproduces an inverse association between smoking and PD, which has been identified in numerous studies^2^. The mechanisms underlying this association are unknown. Though there is ample evidence for a protective role of nicotine in animal models of PD, nicotine has failed in clinical trials^8^. Interestingly, in our data, smoking was not correlated with asthma/COPD but was rather part of the cardiovascular cluster 3 and correlated with baby aspirin, atorvastatin, hyperlipidemia, and heart failure. Thus, the common assumption that COPD/asthma is a generally useful marker for smoking is not reflected in our data. Cardiovascular disease has been associated with development of PD and with increased risk of progression,^10^ and these results highlight the need for epidemiologic studies to consider complex interactions and links between cardiovascular traits, cardiovascular medications, and smoking.

Our study also supports previously reported associations between head trauma and pesticide exposure and PD. The evidence for a positive association between head trauma and PD has been mixed. While several studies including a 2013 meta-analysis have demonstrated an association^11^, large population level Scandinavian studies have not. Recall bias and the timing of injury are potential complicating factors. Similarly, while several large epidemiologic studies have linked pesticide exposure to PD risk, in most studies exposure is self-reported, and the role of individual pesticides is unclear.^2^

Our analysis has limitations. For each logistic regression, we excluded subjects who were missing data for any of the 64 variables, rather than only excluding subjects missing the variable in question. This reduced the size of the available cohort and decreased the power of the analysis, but the alternative approach of excluding only subjects missing the variable in question would also have the disadvantage that each analysis would have used a different sample. Further, we considered only a binary yes/no for each exposure, considering the large number of variables analyzed. This highlights the need for future studies examining variables quantitatively rather than categorically. Further, although the HBS questionnaire is extensive, it is not exhaustive. Some medications such as asthma inhalers were not recorded in HBS until recently. Thus, we could not evaluate for associations between asthma medications and PD, which have been identified in recent studies^12^.

Our logistic regression models were adjusted for age and sex only. We did not additionally adjust for other clinical variables, such as smoking or cardiovascular comorbidities, because many of these are themselves part of the 64-variable exposure set under study and are correlated with other exposures within the same analysis (Figure 2A-B). Adjusting each model for variables that lie on the same causal pathway or share correlation structure with the exposure of interest risks overadjustment bias, potentially attenuating or distorting the true association rather than isolating it. Similarly, disease-related characteristics such as UPDRS score and disease duration are only defined for PD cases and cannot be incorporated as covariates in a case-control model predicting diagnostic status across the full cohort. We acknowledge, however, that residual confounding from unmeasured or unadjusted variables remains possible, and that our minimal adjustment strategy, while intended to avoid overadjustment, cannot fully rule out confounding by factors such as education or other socioeconomic variables not systematically captured in this analysis.

We have focused our discussion on the variables that were statistically significant after adjusting for sex and age and after correcting for multiple tests. Beyond these, we found some variables that were nominally statistically significant but did not survive correction for multiple testing (Supplemental Table 2). These results should be interpreted with caution and will require confirmation in other patient cohorts. Discordant findings may be due in part to suppression effects and residual confounding, in which positively or negatively correlated variables (Figure 2) may suppress or inflate each other’s true relation to PD.

In summary, here we provide an exploratory clinical characterization of PD patients in the HBS. Our results confirm some previously reported associations as well as highlight other novel associations. Many of the health variables we have examined here are modifiable, meaning that these results may someday have implications for personalized medicine. Future work will require mechanistic studies to identify gene-environment interactions, to determine which factors are truly causative, and to discover whether modifying them has a neuroprotective or symptomatic benefit.

## Abbreviations

PD: Parkinson’s disease
HBS: Harvard Biomarkers Study

## Acknowledgements

The Harvard Biomarkers Study (HBS) https://amp-pd.org/unified-cohorts/hbs was founded and co-directed by Dr. Clemens Scherzer and Dr. Bradley T. Hyman. After his move to Yale, Dr. Scherzer is directing its expansion, the Yale Harvard Biomarkers Study (YHBS). HBS was seeded by the Harvard NeuroDiscovery Center, with contributions from APDA Center for Advanced Research awards (to C.R.S.), the Michael J Fox Foundation (to C.R.S.), NINDS U01NS082157 (to C.R.S.), U01NS100603 (to C.R.S.), and the Massachusetts Alzheimer’s Disease Research Center NIA P50AG005134.

C.R.S.’s work is supported by NIH grants U01NS095736, U01NS100603, R01AG057331, and R01NS115144, and the American Parkinson Disease Association Center for Advanced Parkinson Research at BWH and Yale.

The study was in part supported by the APDA Center and joint efforts of The Michael J. Fox Foundation for Parkinson’s Research (MJFF) and the Aligning Science Across Parkinson’s (ASAP) initiative. MJFF administers the grant [ASAP-000301] on behalf of ASAP and itself.

## Author Contributions

ALO: manuscript writing, data analysis, interpretation of results. JJL, TR: statistical consultation, manuscript revision, interpretation of results. CRS: HBS design, project design, manuscript revision, interpretation of results. YK, PK, TY, EA, IT, NL: clinical data management and sample collection. AV, MTH, GPH, JP, VK, TMH, BH, DS, JHG, STG, MAS, AYH, A-M W: referring patients, manuscript review.

## Data availability

The data that support the findings of this study are available from the corresponding author upon request. The supplemental files associated with this manuscript have been deposited in Zenodo: https://doi.org/10.5281/zenodo.19583021. The data, code, protocols, and key lab materials used and generated in this study are listed in a Key Resource Table alongside their persistent identifiers at https://doi.org/10.5281/zenodo.19924992.

An earlier version of this manuscript was posted to medrxiv on February 1, 2022 at doi:https://doi.org/10.1101/2022.02.01.22270276. The most recent version was posted to medrxiv on May 4, 2026. The code used in this manuscript is available at https://github.com/abbyolsen/HBS.

## Supplemental Figure Legends

**Supplemental Figure 1:** Predicted probability of being in the PD group by sex and age for each PD-associated variable.

**Supplemental Figure 2:** High prevalence of sleep and mood disorders in PD patients. A. PD patients are more likely than healthy controls to have multiple sleep and mood disorders. B. Among PD patients with multiple sleep and mood disorders, a large majority have depression.

**Supplemental Figure 3:** Schematic demonstrating the variables examined in this study. Variables span domains of past medical history, medication and supplement use, social history, and environmental exposures. They capture functions of many organ systems. Variables that were significant in the logistic regression are indicated with *.

## Notes

### Competing Interest Statement

C.R. Scherzer is named as co-inventor on a US patent application on sphingolipids biomarkers that is jointly held by Brigham & Womens Hospital and Sanofi. CRS has consulted for Sanofi Inc. and Calico; has collaborated with Pfizer, Opko, and Proteome Sciences, Genzyme Inc., and Lysosomal Therapies; is on the Scientific Advisory Board of the American Parkinson Disease Association; has served as Advisor to the Michael J. Fox Foundation, NIH, Department of Defense, and Google; has received funding from the NIH, the U.S. Department of Defense, the Michael J. Fox Foundation, and the American Parkinson Disease Association. A-M. Wills reports research funding from the Parkinsons Foundation, has participated in clinical trials funded by Sanofi/Genzyme, Roche, Transposon therapeutics and received consultant payments from Accordant, CVS/Caremark, and Sanofi/Genzyme. A.L. Olsen, J.J. Locascio, I. Tuncali, N. Laroussi, E. Abatzis, P. Kamenskaya, Y. Kuras, T. Yi, A. Videnovic, M.T. Hayes, G.P.H. Ho, J. Paulson, V. Khurana, T.M. Herrington, B.T. Hyman, D.J. Selkoe, J.H. Growdon, S.N. Gomperts, T. Riise, M.A. Schwarzschild, and A.Y. Hung report no disclosures relevant to the manuscript.

### Author Declarations

The IRB of Mass General Brigham gave ethical approval of this work.

### Summary of Updates

- Data availability statement has been updated with correct zenodo links - License changed to CCBY

