## Supplemental Table 1 for "Health phenome of Parkinson’s patients reveals prominent mood-sleep cluster"

Supplemental Table 1: The Harvard Biomarkers Study Questionnaire

| | |
|---|---|
| Supplemental Table 1 | |
| **Environmental Exposure Questionnaire in Harvard Biomarkers Study** | |
| **Past Medical History** | |
| Have you been diagnosed with any of the following medical conditions? Please circle either YES or NO for each of the medical conditions? | |
| | Depression active within the past two years? |
| | Depression prior to two years? |
| | Anxiety Disorder? |
| | Sleep apnea? |
| | Periodic limb movements of sleep (PLMS)? |
| | REM behavior disorder (RBD)? |
| | Mild cognitive impairment (MCI)? |
| | High blood pressure? |
| | High cholesterol? |
| | Diabetes mellitus? |
| | Osteoarthritis? |
| | Rheumatoid arthritis? |
| | Coronary artery disease? |
| | Congestive heart failure? |
| | Hypothyroidism? |
| | Asthma/COPD? |
| | Restless leg syndrome? |
| | Do you have any other medical condition or disease? If YES, please list other medical conditions: |
| **Concomitant Medications** | |
| Do you use any of the following medications on a regular basis (at least two times per week)? Circle YES or NO for each medication. | |
| | Tylenol (acetaminophen) |
| | Aspirin or aspirin-containing products (325 mg/tablet or more) |

| | |
|---|---|
| | Atenolol |
| | “Baby” or low dose aspirin (81 mg/tablet or less) |
| | Celebrex (celecoxib) |
| | Cozaar (losartan, hydrochlorothiazide losartan/potassium) |
| | Crestor (rosuvastatin) |
| | Cymbalta (duloxetine) |
| | HCTZ (hydrochlorothiazide) |
| | Advil, Motrin, Nuprin (ibuprofen) |
| | Lescol (fluvostatin) |
| | Meavacor (lovastatin) |
| | Anaprox, Aleve, Naprelan, Naprosyn (naproxen) |
| | Pepcid (famotidine) |
| | Pravachol (pravastatin) |
| | Prevacid (lansoprazole) |
| | Prilosec (omeprazole) |
| | Protonix (pantoprazole) |
| | Vytorin (ezetimibe/simvastatin) |
| | Wellbutrin (buproprion) |
| | Xanax (alprazolam) |
| | Zestril (lisinopril) |
| | Zetia (ezetimibe) |
| | Zocor (simvastatin) |
| | Zoloft (sertraline) |
| | Do you use any other medications on a regular basis? If YES, please specify: |
| **Nutritional Supplements** | |
| Do you take nutritional supplements? If YES, please specify: | |
| | Coenzyme Q10? If YES, please specify dosage per day: |
| | Creatine? If YES, please specify dosage per day: |
| | Multivitamin supplements? |
| | Vitamin E supplements? |
| | Calcium? |
| | Zinc? |

| | |
|---|---|
| | Glucosamine/chondroitin? |
| | Vitamin C? |
| | Vitamin D? |
| **Parkinson's Disease Risk Factors** | |
| | Have you ever had any major head trauma? |
| | If YES, did you lose consciousness? |
| | Have you ever taken a neuroleptic or antipsychotic medication? |
| | If YES, please specify drug and status: |
| | Haloperidol (Haldol) Currently taking: No longer taking: |
| | Risperdal Currently taking: No longer taking: |
| | Mellaril Currently taking: No longer taking: |
| | Stelazine Currently taking: No longer taking: |
| | Other: Currently taking: No longer taking: |
| | Have you ever taken any of the following medications? |
| | If YES, please specify drug and status: |
| | Metoclopramide (reglan) Currently taking: No longer taking: |
| | Compazine Currently taking: No longer taking: |
| | Phenergan Currently taking: No longer taking: |
| | Amiodarone Currently taking: No longer taking: |
| | Have you ever had a brain infection (encephalitis)? |
| | Have you ever been knowingly exposed to pesticides or herbicides? |
| | If YES, which of the following best describes the type and frequency of exposure (check all those that apply): |
| | ☐ Single event (i.e. accident)<br>Details: Year if known, Name of chemical if known |
| | ☐ Continuous exposure for personal use in garden or household<br>Details: Average frequency (once a year or less, 2-5 times/year, 6-11 times/year, monthly or more frequency), Period of exposure, Name of chemicals if known |
| | ☐ Continuous occupational exposure in farming/agriculture<br>Details: Average frequency (once a year or less, 2-5 times/year, 6-11 times/year, monthly or more frequency), Period of exposure, Name of chemicals if known |
| | ☐ Continuous occupational exposure in manufacturing<br>Details: Average frequency (once a year or less, 2-5 times/year, 6-11 times/year, monthly or more frequency), Period of exposure, Name of chemicals if known |
| | ☐ Continuous occupational exposure due to proximity to sprayed area |

| | |
|---|---|
| | Details: Average frequency (once a year or less, 2-5 times/year, 6-11 times/year, monthly or more frequency), Period of exposure, Name of chemicals if known |
| | ☐ Other type of exposure<br>Details: Average frequency (once a year or less, 2-5 times/year, 6-11 times/year, monthly or more frequency), Period of exposure, Name of chemicals if known |
| | Have you ever had heavy metal poisoning? If YES, briefly explain the poisoning: |
| | Have you ever worked in any of the following professions? If YES, please check all applicable professions: |
| | ☐ Welding |
| | ☐ Metal melting |
| | ☐ Metal purification |
| | ☐ Galvanization |
| | ☐ Milling |
| | ☐ Petrochemistry |
| | ☐ Agriculture |
| | ☐ Wood processing |
| | ☐ Textile or industrial painting |
| | Did you ever regularly drink water from a well? If Yes, what time period did you drink well water? |
| | Are you a vegetarian or vegan? |
| **Social History** | |
| | Do you drink caffeinated coffee (not decaf)? |
| | If YES, how many cups per day on average? |
| | Has your coffee consumption changed over the past 10 years? |
| | If YES, has there been a general increase or general decrease in your consumption over a 10 year period? |
| | Do you drink tea (not decaffeinated? |
| | If YES, how many cups per day on average? |
| | Has your tea consumption changed over the past 10 years? |
| | If YES, has there been a general increase or general decrease in your consumption over a 10 year period? |
| | Do you drink caffeinated soda? |
| | If YES, how many ounces per day on average? |
| | Has your soda consumption changed over the past 10 years? |
| | If YES, has there been a general increase or general decrease in your consumption over a 10 year period? |
| | Do you drink alcohol? |
| | If YES, how many drinks do you have on an average day? |

| | |
|---|---|
| | Have you consumed alcohol heavily in the past? |
| | If YES, for how many years? |
| | Has your alcohol consumption changed over the past 10 years? |
| | If YES, has there been a general increase or general decrease in your consumption over a 10 year period? |
| | Do you or have you ever smoked cigarettes, cigars, or pipes, at least once a day for a year's time? |
| | If YES, do you currently smoked or have you quit smoking cigarettes, cigars, or pipes? |
| | ☐ Currently smoke, please fill in the chart<br>Details: Average number smoked per day (cigarettes, cigars, pipes); Age began smoking |
| | ☐ If you have quit smoking cigarettes, cigars, or pipes, please fill in the chart<br>Details: Average number smoked per day (cigarettes, cigars, pipes); Age began smoking, Age quit |
| | Whether or not you smoke, how many hours a day are you exposed to the cigarette smoke of others on average? |
