## Supplemental Table 2 for "Health phenome of Parkinson’s patients reveals prominent mood-sleep cluster"

| Class | Variable | p-value, age/sex adjusted | FDR (age/sex adjusted) | Odds Ratio of being PD |
|---|---|---|---|---|
| Exposures | Alcohol | 0.1057 | 0.3382 | 0.79 |
| Exposures | Antiemetic use | 0.0396 | 0.181 | 3.09 |
| Exposures | Coffee | 0.1593 | 0.4433 | 0.8 |
| Exposures | Encephalitis | 0.7605 | 0.9293 | 1.29 |
| Exposures | Head trauma | 0.0002 | 0.0021 | 2.33 |
| Exposures | Industrial profession | 0.2917 | 0.6438 | 0.79 |
| Exposures | Metal poisoning | 0.7564 | 0.9293 | 1.23 |
| Exposures | Neuroleptic use | 0.9133 | 0.9823 | 0.95 |
| Exposures | Pesticide exposure | 0.0001 | 0.0013 | 1.87 |
| Exposures | Smoking | <0.0001 | 0.0013 | 0.57 |
| Exposures | Soda | 0.5048 | 0.788 | 1.11 |
| Exposures | Tea | 0.9025 | 0.9823 | 0.98 |
| Exposures | Vegetarian | 0.4707 | 0.7828 | 1.46 |
| Meds | Acetaminophen | 0.5478 | 0.8153 | 1.15 |
| Meds | Alprazolam | 0.3386 | 0.6519 | 2.8 |
| Meds | Aspirin (325 mg) | 0.6669 | 0.9081 | 0.9 |
| Meds | Atenolol | 0.7841 | 0.9293 | 1.11 |
| Meds | Atorvastatin | 0.064 | 0.2731 | 0.69 |
| Meds | Baby aspirin (81 mg) | 0.9125 | 0.9823 | 0.98 |
| Meds | Buproprion | 0.0224 | 0.1103 | 10.28 |
| Meds | Celecoxib | 0.9085 | 0.9823 | 0.94 |
| Meds | Duloxetine | 0.2228 | 0.5704 | 3.64 |
| Meds | Ezetimibe | 0.0151 | 0.0827 | 0.13 |
| Meds | Ezetimibe/Simvastatin | 0.9819 | 0.9823 | Incalcuably large |
| Meds | Famotidine | 0.6195 | 0.8895 | 0.7 |
| Meds | Fluvastatin | 0.9801 | 0.9823 | Incalcuably large |
| Meds | Hydrochlorothiazide (HCTZ) | 0.3021 | 0.6445 | 0.73 |
| Meds | Hypertension (HTN) | 0.9583 | 0.9823 | 0.99 |
| Meds | Ibuprofen | 0.0145 | 0.0827 | 1.74 |
| Meds | Lansoprazole | 0.3262 | 0.6519 | 0.47 |
| Meds | Lisinopril | 0.7213 | 0.9293 | 1.11 |
| Meds | Losartan | 0.1523 | 0.4431 | 4.42 |
| Meds | Lovastatin | 0.4515 | 0.7828 | 1.8 |
| Meds | Naproxen | 0.2739 | 0.6261 | 0.67 |
| Meds | Omeprazole | 0.8702 | 0.9823 | 0.96 |
| Meds | Pantoprazole | 0.4667 | 0.7828 | 1.76 |
| Meds | Pravastatin | 0.2725 | 0.6261 | 1.73 |
| Meds | Rosuvastatin | 0.7808 | 0.9293 | 0.88 |

| | | | | |
|---|---|---|---|---|
| Meds | Sertraline | 0.1057 | 0.3382 | 2.78 |
| Meds | Simvastatin | 0.6526 | 0.908 | 1.12 |
| PMH | Anxiety | <0.0001 | 0.0013 | 3.31 |
| PMH | Asthma/Chronic obstructive pulmonary disease (COPD) | 0.477 | 0.7828 | 0.83 |
| PMH | Coronary artery disease | 0.1805 | 0.4813 | 1.65 |
| PMH | Depression | <0.0001 | 0.0013 | 3.11 |
| PMH | Diabetes Mellitus (DM) | 0.125 | 0.381 | 1.69 |
| PMH | Heart failure | 0.4933 | 0.788 | 0.63 |
| PMH | Hyperlipidemia (HLD) | 0.6254 | 0.8895 | 1.08 |
| PMH | Hypothyroidism | 0.2639 | 0.6261 | 1.32 |
| PMH | Mild cognitive impairment (MCI) | 0.0972 | 0.3382 | 5.52 |
| PMH | Osteoarthritis (OA) | 0.544 | 0.8153 | 1.13 |
| PMH | Periodic limb movements of sleep (PLMS) | 0.3411 | 0.6519 | 1.69 |
| PMH | REM behavior disorder (RBD) | 0.9809 | 0.9823 | Incalcuably large |
| PMH | Restless leg syndrome (RLS) | 0.0027 | 0.0192 | 4.12 |
| PMH | Rheumatoid arthritis (RA) | 0.7125 | 0.9293 | 1.17 |
| PMH | Sleep apnea | 0.0019 | 0.0152 | 2.58 |
| Vitamins | Calcium | 0.3565 | 0.6519 | 0.8 |
| Vitamins | Coenzyme-Q | <0.0001 | 0.0013 | 2.98 |
| Vitamins | Creatine | 0.9823 | 0.9823 | Incalcuably large |
| Vitamins | Glucosamine | 0.7744 | 0.9293 | 1.12 |
| Vitamins | Multivitamin | 0.0155 | 0.0827 | 0.71 |
| Vitamins | Vitamin C | 0.0891 | 0.3354 | 1.73 |
| Vitamins | Vitamin D | 0.0007 | 0.0064 | 2.18 |
| Vitamins | Vitamin E | 0.0783 | 0.3132 | 0.68 |
| Vitamins | Zinc | 0.3528 | 0.6519 | 0.65 |

| Confidence Interval |
|---|
| 0.06-1.05 |
| 1.17-10.67 |
| 0.58-1.09 |
| 0.3-8.94 |
| 1.51-3.73 |
| 0.52-1.23 |
| 0.36-5.65 |
| 0.39-2.67 |
| 1.37-2.6 |
| 0.43-0.75 |
| 0.82-1.5 |
| 0.74-1.31 |
| 0.56-4.53 |
| 0.72-1.91 |
| 0.5-52.77 |
| 0.57-1.47 |
| 0.54-2.53 |
| 0.47-1.03 |
| 0.73-1.34 |
| 2.18-183.84 |
| 0.32-3.39 |
| 0.68-67.66 |
| 0.023-0.731 |
| 1.88- |
| 0.19-3.36 |
| 0.46- |
| 0.41-1.36 |
| 0.74-1.33 |
| 1.13-2.77 |
| 0.11-2.42 |
| 0.63-2.06 |
| 0.88-80.42 |
| 0.47-11.86 |
| 0.33-1.42 |
| 0.56-1.7 |
| 0.47-11.5 |
| 0.71-5.17 |
| 0.38-2.3 |

| |
|---|
| 0.93-11.99 |
| 0.7-1.87 |
| 2.01-5.75 |
| 0.5-1.42 |
| 0.83-3.63 |
| 2.1-4.71 |
| 0.9-3.46 |
| 0.18-2.90 |
| 0.8-1.46 |
| 0.82-2.2 |
| 1.13-99.58 |
| 0.78-1.66 |
| 0.64-5.8 |
| 5.86- |
| 1.81-12.1 |
| 0.53-2.97 |
| 1.47-4.92 |
| 0.5-1.29 |
| 1.89-4.92 |
| 2.41- |
| 0.54-2.58 |
| 0.54-0.94 |
| 0.95-3.4 |
| 1.4-3.45 |
| 0.44-1.06 |
| 0.27-1.73 |
