## Supplementary figures and images for "Health phenome of Parkinson’s patients reveals prominent mood-sleep cluster"

### Supplemental Figure 1

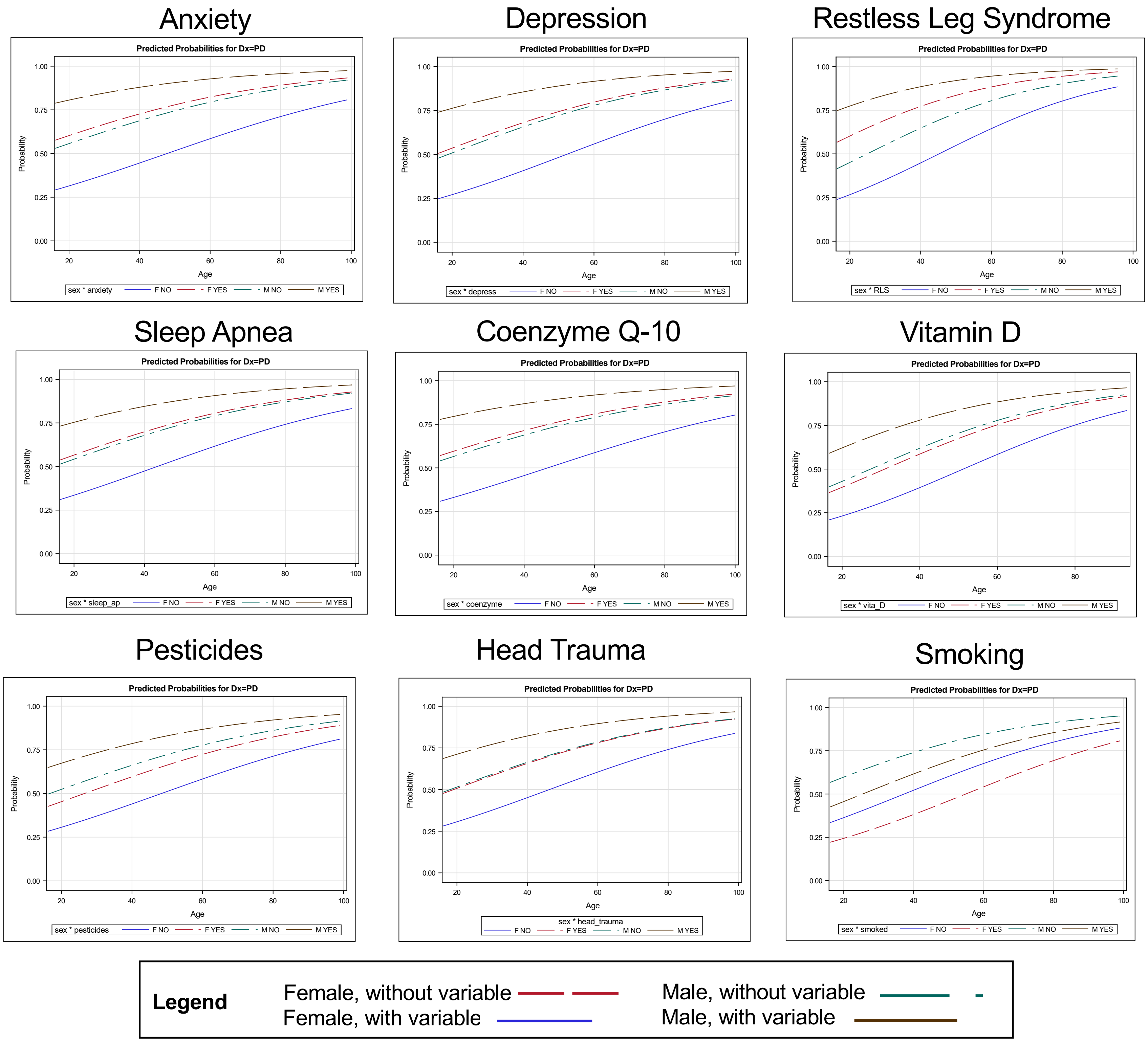

### Supplemental Figure 2

#### Supplemental Figure 2

**A**

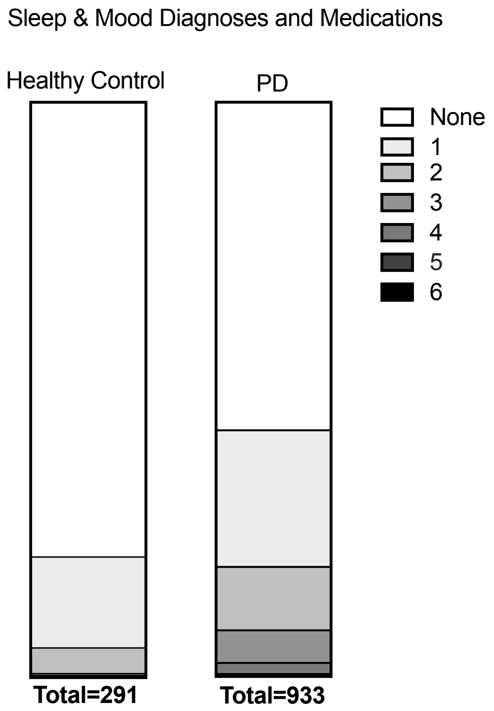


**B**

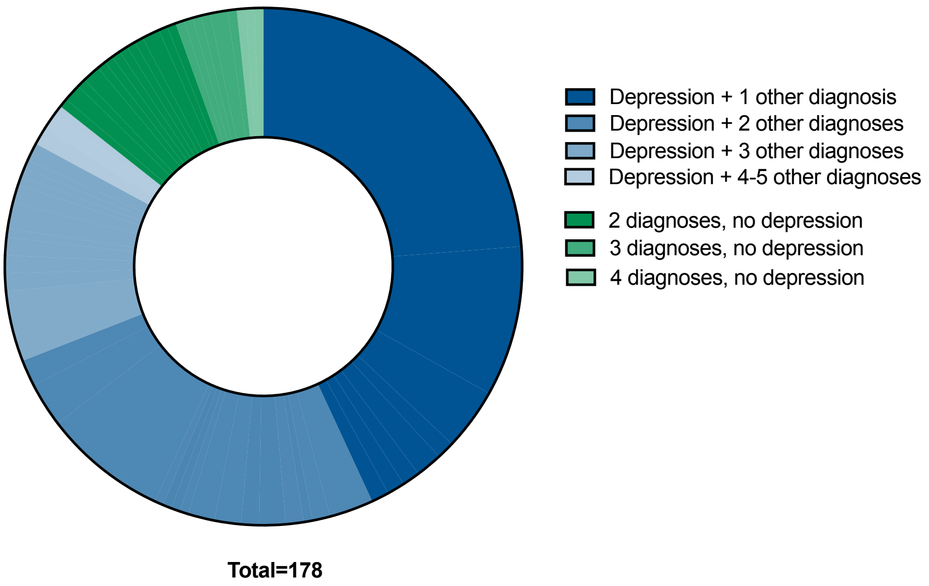

### Supplemental Figure 3

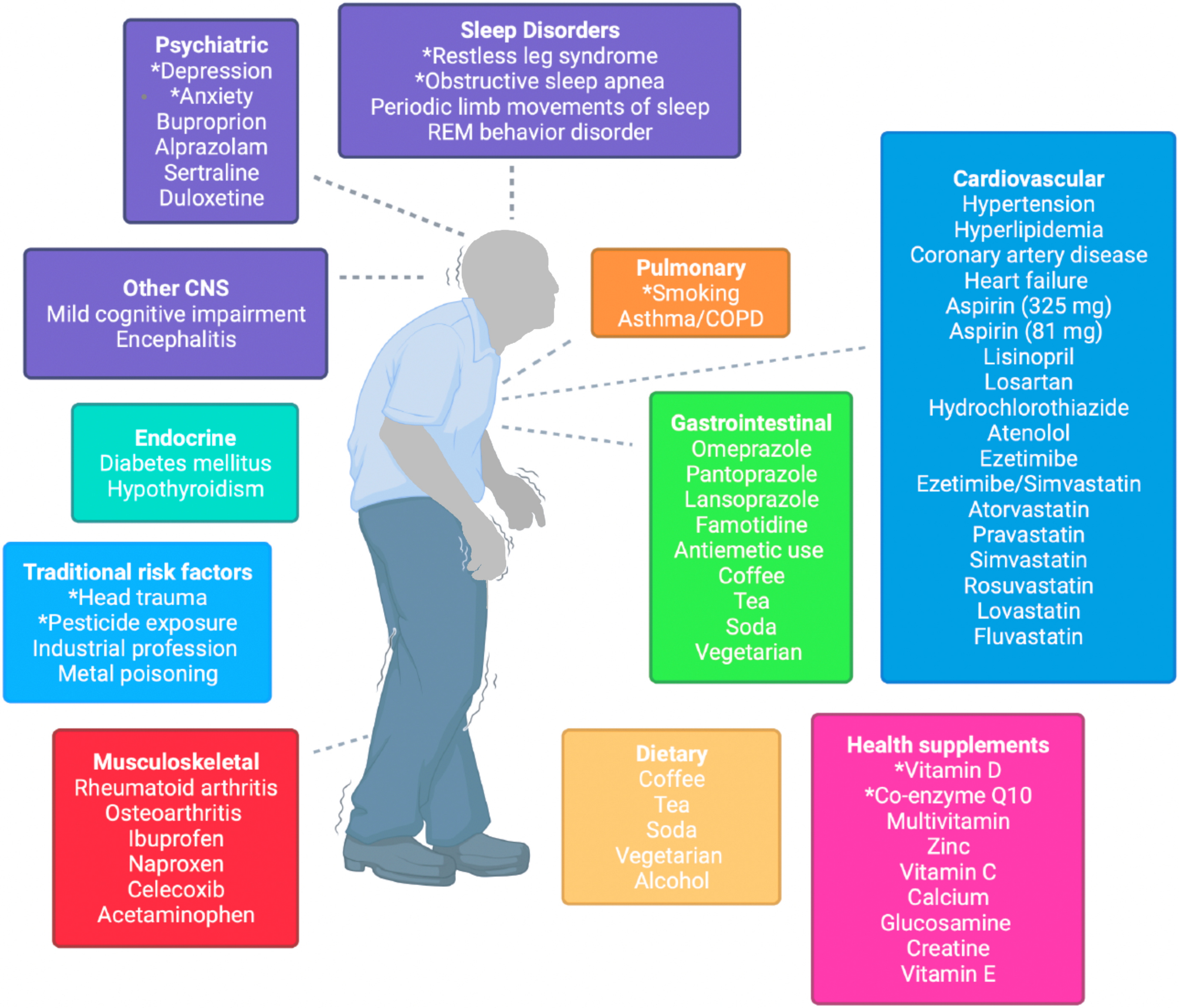
